# Multi-region Multiomic Random Forest Toxicity Modeling of Radiation Pneumonitis

**DOI:** 10.1101/2024.05.23.24307616

**Authors:** Saurabh S. Nair, Ramon M. Salazar, Alexandra O. Leone, Ting Xu, Zhongxing Liao, Laurence E. Court, Joshua S. Niedzielski

## Abstract

**Purpose:** Radiation pneumonitis (RP) is a major dose-limiting toxicity resulting from non-small-cell lung cancer (NSCLC) radiotherapy. Multiomic features (radiomics and dosiomics) could provide additional predictive information as compared to traditionally used clinical and dose-volume histogram (DVH) parameters. We aimed to investigate the utility of multiomic features to improve RP toxicity models.

**Methods:** Out of 329 NSCLC patients considered, 85 patients (25.84%) were found to have toxicity ≥ grade 2 RP per CTCAE v5.0. A total of 422 radiomic and dosiomic features were extracted. Four toxicity prediction model types were created using clinical factors together with respective features from one of the following groups: (a) DVH (base model), (b) whole lung radiomics and dosiomics (WL-RD), (c) multi-region radiomics and dosiomics (MR - RD) and (d) multi-region DVH, radiomics and dosiomics (MR-DVHRD). Toxicity models were created using a random forest classifier with a Monte Carlo cross-validation approach of 100 iterations, and a training/test split of 80%/20%, respectively. Model predictive performance was evaluated by area under the receiver operating characteristic curve (AUC) and area under the precision-recall curve (AUPRC).

**Results:** The AUC and AUPRC values (mean ± standard deviation) for the 4 model types were 0.81±0.04/0.70±0.06 (base model), 0.82±0.05/0.73±0.08 (WL-RD, p<0.05), 0.83±0.06/0.75±0.08 (MR–RD, p<0.05), and 0.82±0.05/0.72±0.08 (MR-DVHRD, p<0.05), respectively, wherein a paired test compared the performance metrics of omic models with the base model built on each iteration of cross0020validation.

**Conclusions:** All multiomic model types outperformed the base DVH model. MR-RD model had the best performance among all model types.

## Introduction

Lung cancer is the leading cause of cancer-related mortality worldwide.^1^ Non-small-cell lung cancer (NSCLC) accounts for 85% of all lung cancer cases.^2^ Radiation therapy (RT) plays a crucial role in the management of unresectable and locally advanced NSCLC, as RT is the primary treatment modality for inoperable NSCLC. Radiation pneumonitis (RP) remains a dose-limiting toxicity that degrades patient quality-of-life, with symptomatic RP occurring in about 15–40% of lung RT patients.^3^ RP can lead to irreversible fibrosis and dyspnea, with possible mortality in severe cases.^3^ Non-effective therapeutics and persistent RP remains one of the top dose-limiting toxicities that influences RT planning.^4,5^ Various clinical factors like age, gender, tumor location and smoking status have been studied in order to predict RP^6^, but the findings have been inconsistent due to sample size^7^ and the heterogeneity of patient populations.^8^ Metrics derived from dose-volume histograms (DVHs) have also been used to predict the incidence of RP with mean lung dose (MLD) and volume of lung receiving ≥20Gy (V20) being the most common metrics related to RP.^9–12^ However, clinical and DVH feature-based toxicity models have been shown to have suboptimal predictive performance.^13^

Recently, machine learning has emerged in the field of radiation oncology for radiation-induced toxicity outcome prediction modeling (OPM). A random forest machine learning classification approach has been shown to outperform other algorithms for outcome predictions with clinical data among some of the widely used machine learning classification approaches.^14^ A process termed radiomics involves extracting quantitative features from medical images, such as computed tomography (CT) scans, and with the aid of a machine learning approach, these features are used to predict therapeutic responses^15–17^. Recently, dosiomics, which is similar to radiomic analyses, but instead uses 3D dose distributions as the feature space, has been introduced into OPM analyses. These omic features are derived from the extracted radiotherapy dose distributions of patients to predict therapeutic responses. It is possible that dosiomic variables can better predict RP than DVH metrics since they retain the 3D information of the radiation dose distribution.^18,19^. Recent studies have also demonstrated the effectiveness of incorporating additional radiomic features derived from a very small region surrounding the tumor into OPM analyses, resulting in improved prediction accuracy^20–22^. Combining multi-region radiomics and dosiomics features into a multiomic OPM approach, based on shape, statistical, and textural patterns from CT Imaging and 3D dose distributions, could provide additional data that improves outcome prediction models.

The aims of this work were to investigate the utility of radiomic and dosiomic features over traditional DVH features, as well as to examine the impact of using multiomic features from a subregion of normal lung to improve RP OPMs.

## Materials and Methods

### Patient Data

We included 329 NSCLC patients treated at our institution between October 2006 to March 2019 with either intensity modulated radiation therapy/volume-modulated arc therapy (IMRT/VMAT, n=190), passive-scatter proton therapy (PSPT, n=92), or intensity-modulated proton therapy (IMPT, n=47). The CT and dose grid spacing were 2.5 mm and 3×3×3 mm, respectively. All patients received fractionated doses between 1.8–3.0 Gy per fraction for total doses between 60–74 Gy. The study exclusion criteria were: (a) presence of acute lung infection, (b) prior thoracic surgery, (c) patients having <6-month follow-up period unless toxicity noted, (d) patients with a previous history of thoracic radiation therapy. All patients were treated with either induction, concurrent, or adjuvant chemotherapy in addition to RT.

### Toxicity Evaluation

Radiation Pneumonitis was graded from 0 to 5 according to the Common Terminology Criteria for Adverse Events (CTCAE v5.0).^23^ The approximate time for RP to develop in a patient after radiation therapy is between 1–6 months.^24,25^ A patient with an RP grade ≥2 was considered symptomatic for RP and used as the binary endpoint for this study.

### Data Analysis Workflow

The data analysis workflow is illustrated in **Fig 1**. Radiomic and dosiomic features were calculated using CT and dose images, respectively, focusing on two regions of interest (ROI). First was the total lung minus the gross tumor volume (GTV). To study the utility of incorporating additional information about tumor microenvironment in enhancing efficiency of RP OPMs, multomic features were also extracted from a 20mm ring structure created from normal lung surrounding the GTV (termed, “multi-region multiomic features”). Once all features were extracted, we conducted a Spearman correlation analysis to remove heavily correlated features with a correlation coefficient threshold of >0.85 for redundant features^18^. Next, we used a random forest classifier to create predictive toxicity models of 4 different types using the selected feature categories: (a) base DVH model, (b) whole lung based radiomic & dosiomic (WL-RD) model, (c) whole lung and ring (multi-region) based radiomic & dosiomic (MR-RD) model, and (d) multi-region DVH + radiomic + dosiomic (MR-DVHRD) model. The performance metrics between the base and mulitomic models were then evaluated. Further details on each component of the study analysis are presented in the following subsections.

**Figure 1.**
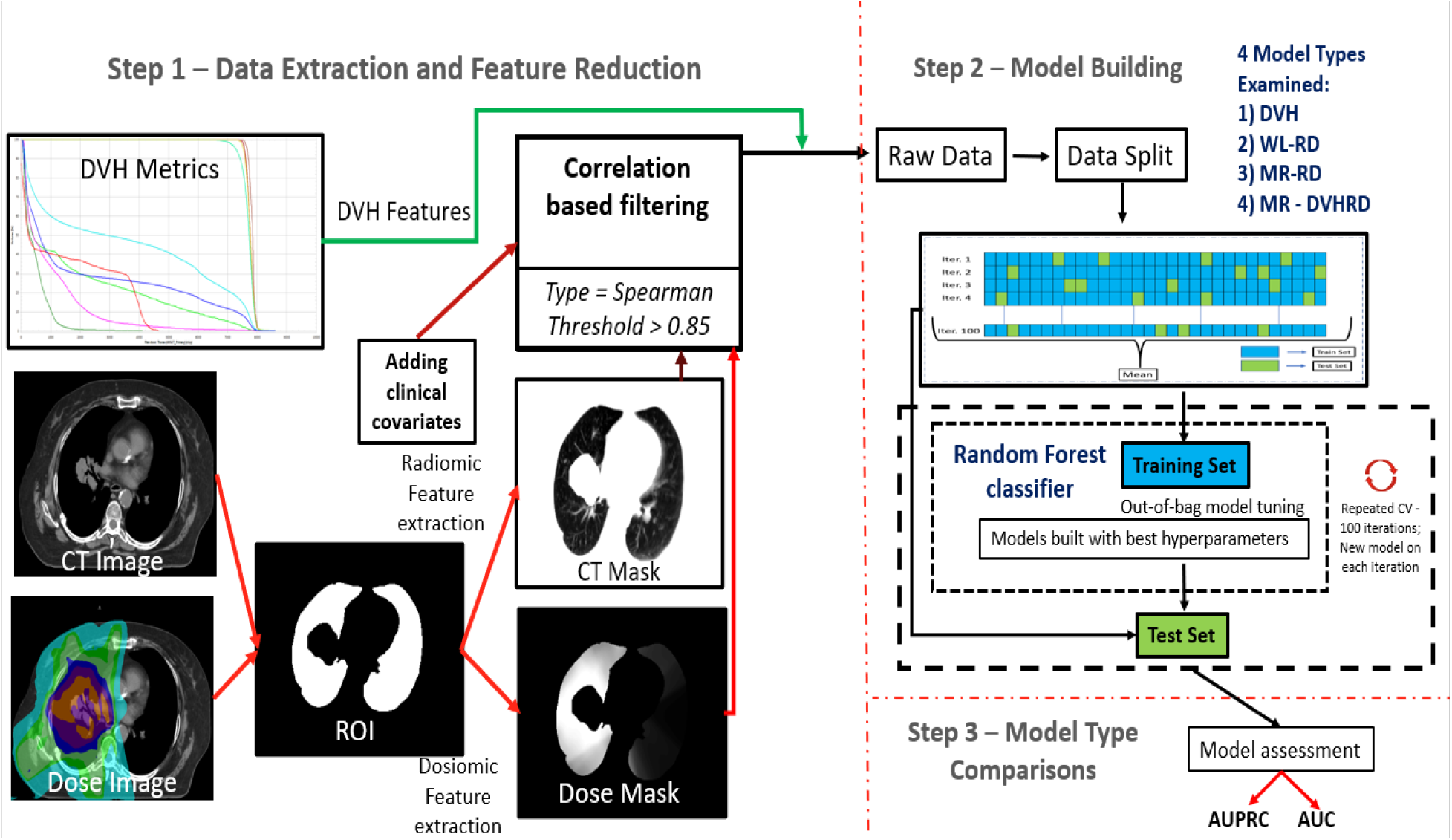
Overall Study Workflow. DVH, radiomic and dosiomic features are extracted, followed by feature reductions for omic features in the first step. 4 OPM types are built and performance metrics between the DVH and omic model types are compared in the second step. *Abbreviations: DVH, dose volume histograms; ROI, region of interest; AUC, area under the curve; AUPRC, area under precision recall curve; CV, cross validation; WL, whole lung; RD, radiomic & dosiomic; MR, multi-region*.

### Feature Extraction and Selection

A total of 422 data features were used for this study (13 clinical, 18 dosimetric, 182 dosiomic and 210 radiomic). The clinical features were gender, age, smoking status, cancer stage, histology, type of chemotherapy, tumor position, gross tumor volume (GTV), and performance status. The description of the clinical features is provided in the **Supplemental Table S1**. The dosimetric, radiomic, and dosiomic features were first extracted from the whole-lung ROI. The dosimetric covariates include mean lung dose, max lung dose, the relative volume receiving at least a given dose (i.e. the value obtained for VxGy[%] refers to the percent volume of a structure receiving at least xGy) from rV5 to rV60 (in increments of 5 Gy). These metrics were extracted from the Raystation treatment planning system (RaySearch Laboratories; Stockholm, Sweden). In order to extract features, the DICOM files were first converted into nearly raw raster data (nrrd) using the 3D Slicer software.^26^ Radiomic and dosiomic features were then extracted based on the Imaging Biomarker Standardization Initiative (IBSI)^27^ using the pretreatment CT and dose image using Pyradiomics (v3.0.1) software. The extracted features included: 14 shape, 18 first-order, and 73 second-order features respectively for both ROI’s.^28^ All mulitomic features are described in the **Supplemental Materials**. For feature selection, a previously reported filter-based correlation approach was implemented.^18^. First, Spearman correlation coefficients (CC) were calculated for all features, with features showing a CC of >0.85 being excluded. The reduction criteria were that if two variables had a higher correlation coefficient than the cutoff, the variable with the highest mean absolute correlation was removed.

### Multi-region multiomics

We assume that the greatest effect for normal tissue toxicities are observed closer to the tumor volume and therefore hypothesize that using a sub-region near the GTV could provide more informative multiomic measures. Unique characteristics captured by these features based on tumor-normal tissue interactions and spatial information could be imperative in enhancing the performance of OPMs. Thus, in this study, in addition to whole-lung multiomic features, we created a 20-mm ring subregion ROI of the normal lung around the GTV and extracted multiomic features to examine the added potential of the features close to the tumor in predicting RP. **Figure 2** shows the sub-region multiomic feature delineation. Once the ring ROI multiomic features are extracted, the data analysis process is the same as depicted in **Fig 1**.

**Figure 2.**
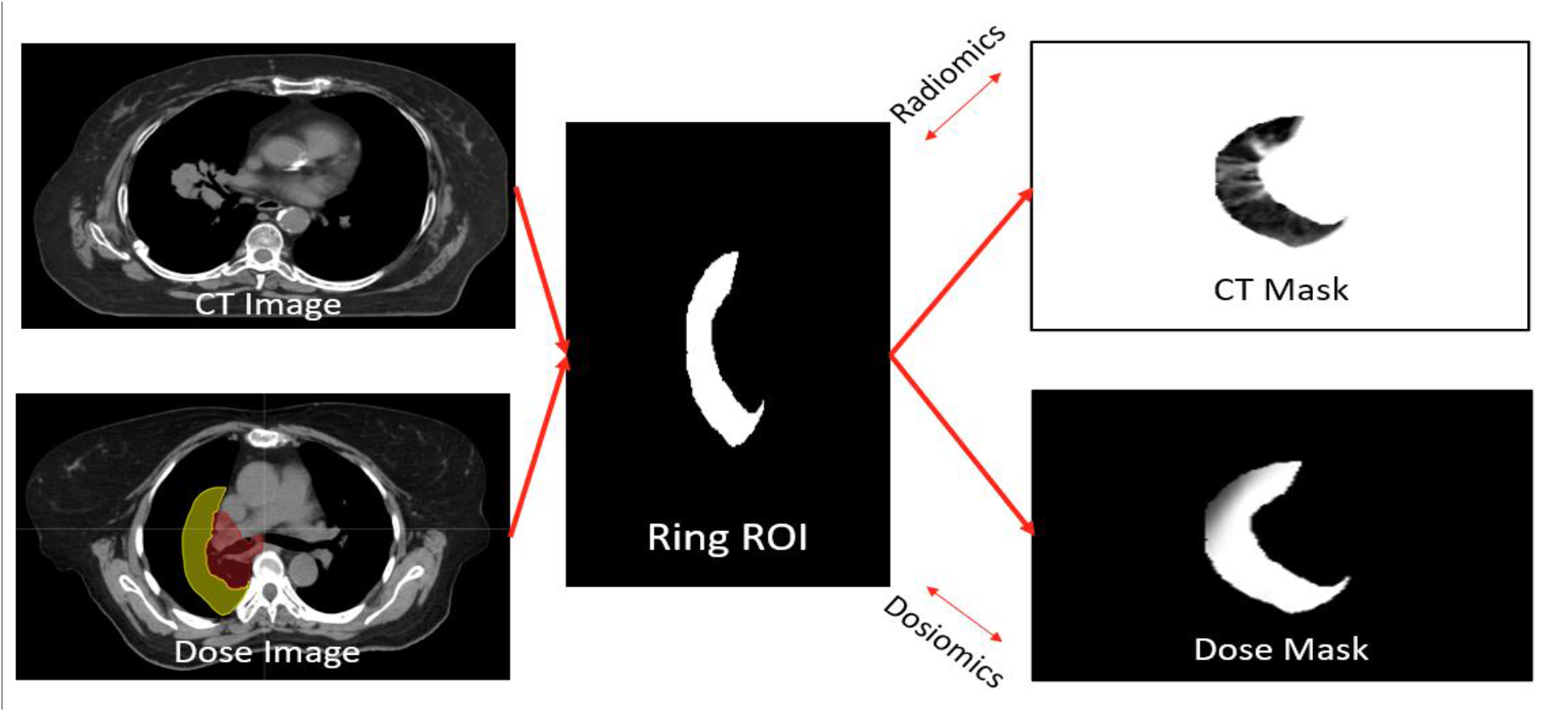
Sub-region (ring) multiomic feature extraction. Radiomic and dosiomic features are extracted for a 20-mm ring around the GTV overlapping the whole-lung ROI, followed by feature reductions and analysis as described in **Fig 1**.

### Model Building

The process of constructing the model and performance metric evaluation is illustrated in **Fig 1 and Fig 2**. For this study, we created 4 toxicity prediction model types using the clinical factors, along with one of the following feature sets: (a) base DVH, (b) whole-lung based radiomic & dosiomic (WL-RD), (c) whole-lung and ring (multi-region) based radiomic & dosiomic (MR-RD), and (d) multi-region DVH + radiomic + dosiomic (MR-DVHRD). The random forest (RF) classification model has been shown to yield the best discriminative performance in radiotherapy outcomes and toxicity predictions and hence has been used in this study.^14^ Hence, to build our toxicity models, we used a random forest classifier. The hyperparameters for the random forest modeling like *mtry* (controls how much randomness is added to the decision tree creation process), *maxnodes* (maximum number of terminal nodes allowed in a tree) and *ntrees* (number of decision trees that are combined to create the final prediction), were set depending on the number of covariates in each of the 4 models.^29^

The toxicity models were created using a repeated cross-validation approach with 100 iterations. This type of cross-validation is a common method used to approximate the generalizability of the modeling process. ^30–32^ Each iteration of cross-validation creates a unique model, with the overall process creating 100 unique models for a given model type. This results in a paired comparison of individual models, where statistical significance of performance metrics can be derived from the distribution of models for each model type. For each iteration, the cohort was randomly split into a training set (80%) and test set (20%). Correlation-based feature reductions were performed for all features on the training set to find out optimal features. The remaining features were then used for RF modeling for the specific model type.

### Model Evaluation

After selecting the optimal hyperparameters using the training set, the performance of the model was tested on the test set. The following metrics were used to assess each model’s performance on the held-out test set: accuracy, area under the receiver operating characteristic curve (AUC), area under the precision-recall curve (AUPRC), precision, and recall. Standard deviation was also computed for performance metric distributions from the repeated cross-validation model building on both the training and test sets. All omic model types were compared to the base model to test the significance of each performance metrics distribution from repeated cross-validation using the paired Wilcoxon sign-rank test, with a p-value of <0.05 being considered as statistically significant. All statistical analyses were performed using R Studio (4.2.1) (Posit, PBC; Boston, Massachusetts). The open-source R package “caret” with randomForest library was used to build all machine learning outcome prediction models.

## Results

### Patient Characteristics and Univariate Analysis

A total of 329 patients were used for this study. The patients who developed a grade 2 or higher RP were 85/329 (25.84% patients positive for RP). The patient characteristics are summarized in **Supplemental Table S1**. The results for the Fisher’s exact test and Mann Whitney U-test for categorial, continuous, and DVH variables are provided in **Supplemental Table S1**.

### Multivariate analysis

The results of all the performance metrics for all models (test set) for predicting RP≥2 is reported in **Table 1**. The results for the training set are reported in **Supplementary Table S3**. All multiomic models outperformed the base model. Also, the MR-RD model type was the best performing model type among all model types. Boxplots comparing AUC and AUPRC for all the 4 model types are also shown in **Figure 2**. These results show that model types with omic features are better than the base model and the MR-RD model type is the best performing model among all models.

**Table 1.** Results of test set evaluation for different model types reported as mean of each metric with one standard deviation as the error. Model type comparison with the base model type (DVH metric) for WL-RD, MR-RD, and MR-DVHRD models is conducted with a one-sided Wilcoxon signed-rank test. Models with multiomic features had a better AUC and AUCPR as compared to the base model with MR-RD model type outperforming all the other model types.

| Model Name | Accuracy | AUC | AUPRC | Precision | Recall |
| --- | --- | --- | --- | --- | --- |
| **Base (DVH)** | 0.86±0.02 | 0.81±0.04 | 0.70±0.06 | 0.74±0.08 | 0.69±0.08 |
| **WL-RD** | 0.86±0.03 | 0.82±0.05* | 0.73±0.08* | 0.73±0.12 | 0.69±0.09 |
| **MR-RD** | 0.86±0.03 | 0.83±0.06** | 0.75±0.08*** | 0.72±0.12 | 0.73±0.10* |
| **MR-DVHRD** | 0.84±0.03 | 0.82±0.05* | 0.72±0.08* | 0.69±0.11 | 0.73±0.11* |

\*p<0.05, \*\*p<0.001, \*\*\*p<0.00001 – Significance level as compared to the base model
*Abbreviations: DVH, dose volume histogram; AUC, area under the curve; AUPRC, area under precision-recall curve; WL, whole lung; RD, radiomic & dosiomic; MR, multi-region.*

The most important (top 10) features are plotted as per rank for the base, WL-RD, MR-RD and MR-DVHRD model types in **Figure 3**. The plots show that for the base model type (**Fig 3A**), the relative V30 and V35 metrics are extremely important followed by relative V25 and mean lung dose. In the WL-RD model type **(Fig. 3B)**, Kurtosis was found to be an important model feature, followed by dosiomic GLSZM Gray Level Variance, and dosiomic GLCM Inverse Variance. In the MR-RD model type **(Fig. 3C)**, Kurtosis was once again shown to be the most important covariate followed by dosiomic GLSZM Gray Level Variance and Interquartile Range. Also, in the MR-DVHRD model type **(Fig. 3D)**, dosiomic GLSZM Gray Level Variance was seen to be the most important covariate followed by rV60, dosiomic GLRLM Gray Level Non-Uniformity Normalized and others. Descriptions for all features are mentioned in the **Supplemental Materials**.

**Figure 3.**
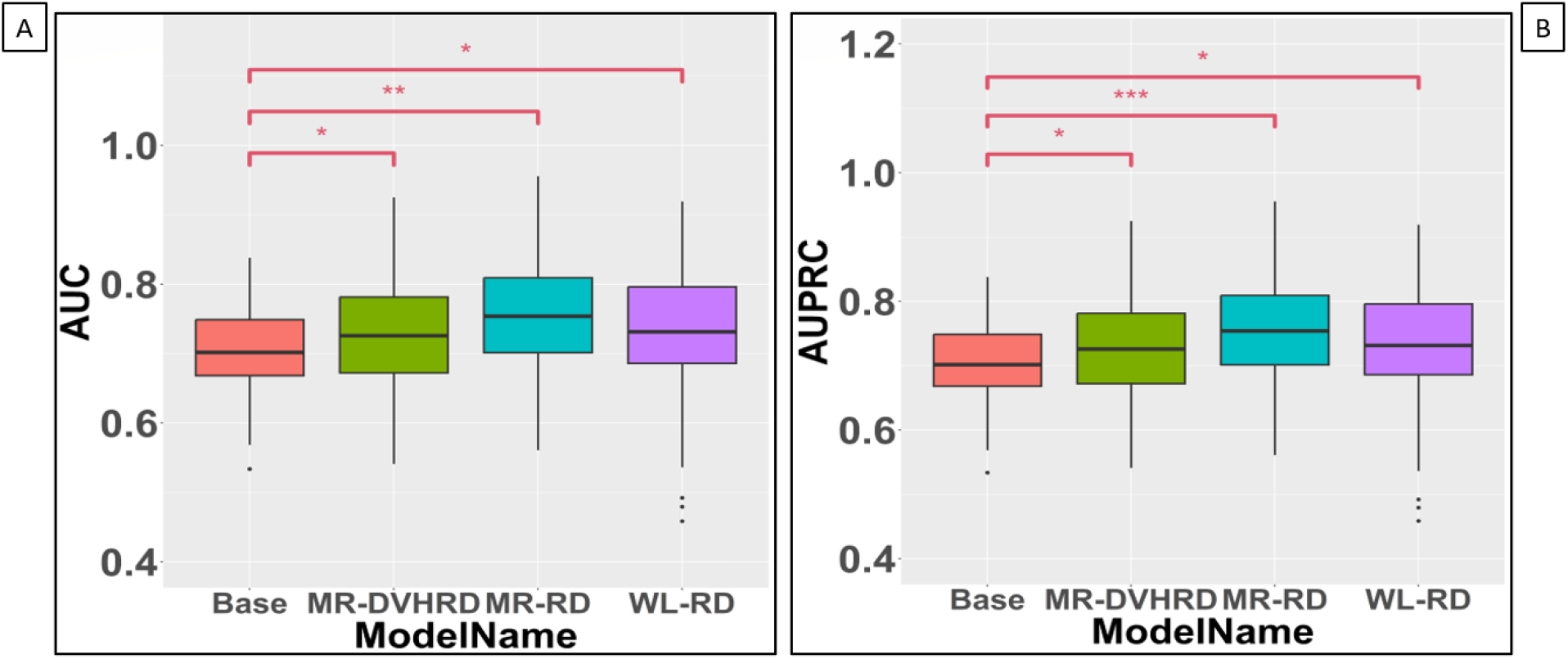
Boxplots comparing performance metrics of all 4 model types. A, Area under the curve (AUC) comparison between model types; B, area under precision-recall curve (AUPRC) comparison between model types. In both comparisons, the performance metrics for the multiomic models are statistically significant as compared to the base model with the MR-RD being the best performing model. A one-sided Wilcoxon ranked significance test was used to check if the distribution of the other models is greater than that of the base model.

**Figure 4.**
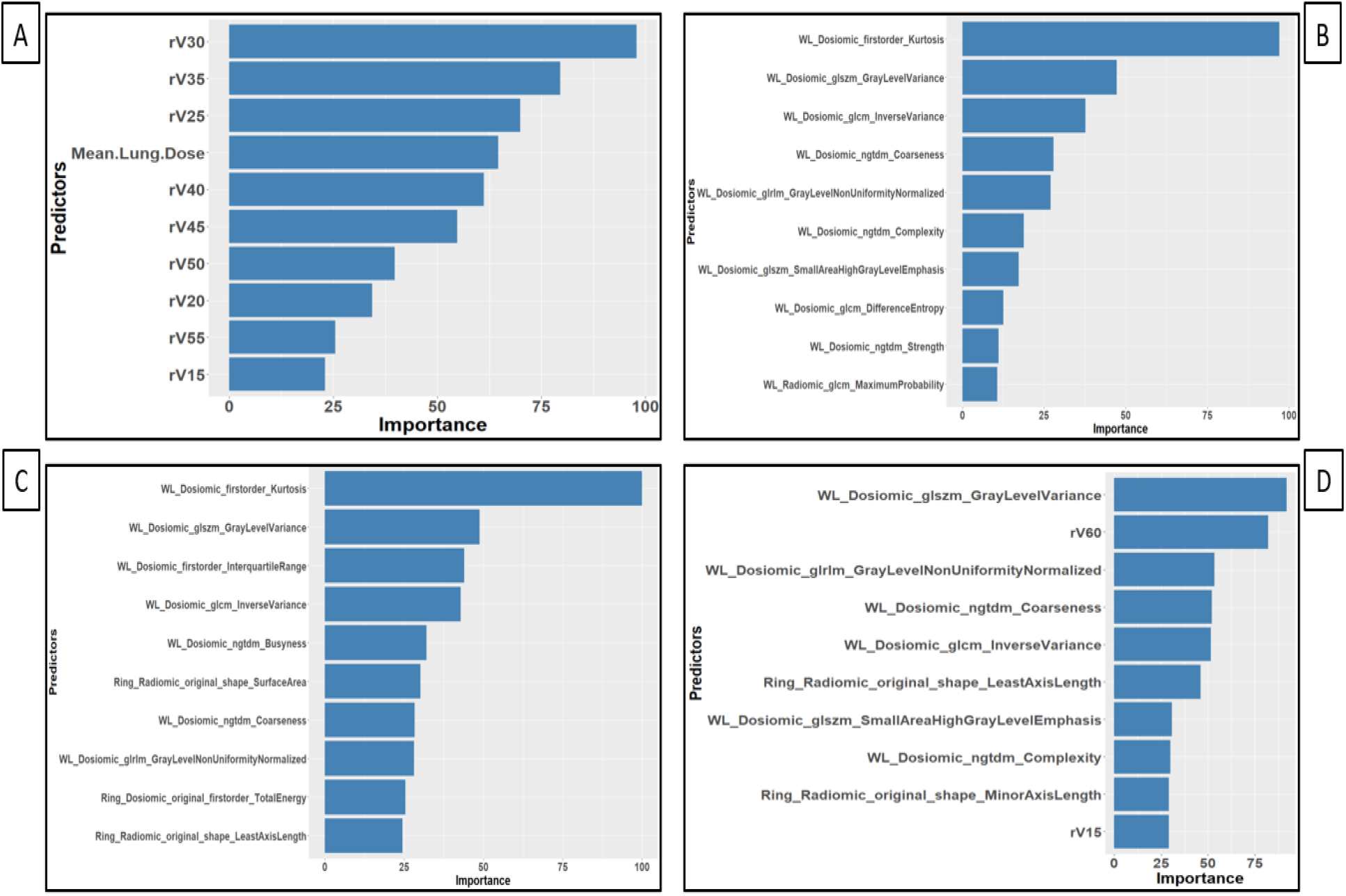
Most Important features for each model type. A, DVH model type top features; B, WL-RD model type top features; C, MR-RD model type top features; D, MR-DVHRD model type top features.

## Discussion

In this study, we compared the performance metrics (AUC, AUPRC, accuracy, precision, and recall) of the base model with the 3 omic model types **(Table 1)** to determine if multiomic features provided improved predictive value for OPMs of RP over using DVH metrics alone. A comparison of the base model with the WL-RD, MR-RD, and MR-DVHRD model types showed that all the multiomic model types outperformed base DVH models. Also, the MR-RD model type had the highest performance of the 4 model types. This shows that multiomic features, especially those extracted from a restricted area surrounding the GTV, has valuable information for predicting RP. To the best of our knowledge, this is the first study to use multi-region multiomic features to predict RP ≥grade 2 in NSCLC patients.

Several studies have analyzed radiomic features from CT for predicting RP. A study by Krafft et al.^33^ demonstrated that when radiomic features were added to clinical and dosimetric features, the resultant model was able to better predict RP signifying the importance of radiomic features. Similarly, another study by Du et al.^34^ used a nomogram-based radiomics approach with esophageal cancer patients and found that adding radiomic features to clinical and DVH metrics enhances the model capability for predicting RP. Dosiomics has been used in various studies to predict RT related treatment efficacy^35^ and prognosis^36^ but few toxicity prediction-based studies exist. A study conducted by Liang et al.^19^ showed that the predictive power of dosiomic features were superior to DVH features; this study indicated that dosiomic features could possess more useful dose information than DVH metrics for predicting toxicity. A combined radiomic and dosiomic study by Zhang et al.^37^ used logistic regression based risk scores to predict RP using clinical, DVH, radiomic and dosiomic features. They found that the prediction model had the best performance when combining, clinical, radiomic, and dosiomic features. However, the paper does not mention which feature set plays the most important role in enhancing model efficacy. In our study as well, the combined clinical, radiomic and dosiomic whole lung model outperformed the base DVH model. However, in our case, the performance of the combined radiomic+dosiomic model type relied mostly on the contributions of the dosiomic features in the model (as illustrated by the top 10 most important features in **Fig. 3B**).

Prior studies have also shown the utility of incorporating additional radiomic features derived from the tumor’s outer region, resulting in improved prediction accuracy^20,21^. A study by Kawahara et al.^22^ also used multi-region radiomic features to predict RP in NSCLC. However, this study did not include two very important aspects: clinicopathologic factors and dosiomic features which have been proven to be vital in predicting RP. In our study, we built two multiomic multi-region model types: (a) MR-RD (including clinical, and multi-region radiomic and dosiomic features) and (b) MR-DVHRD (including DVH, clinical, and multi-region radiomic and dosiomic features), and both these models outperformed the base model. The MR-RD was found to be the best model type among all models. This shows that additional 3-dimensional dose information especially from a finite area surrounding the GTV in terms of ROI geometry, first-order statistics (individual voxel based) and second-order statistics (based on relationship between voxels) have a lot more predictive power as conventional DVH metrics. Features extracted from a sub-region around GTV allows for a more nuanced assessment of normal tissue radiosensitivity and our results suggests that is vital to take these aspects into consideration in the OPMs while predicting RP. This does confirm our hypothesis that incorporating multi-region features into OPMs can identify patients who are at a greater risk of developing RP ≥grade 2, thus mitigating potential adverse effects for future patients. The best performing model types based on our results could be used to run toxicity predictions once the treatment is planned for a specific patient. This would provide the physician with a data-driven prediction of the patient developing grade 2 or above RP before the treatment begins, allowing the physician either to amend the treatment plan or to discuss post-treatment RP risks more accurately with the patient.

The figures for the top 10 most important features, as per Gini index, for the base, WL-RD, ML-RD, and MR-DVHRD model types are shown in **Fig 3**. In the WL-RD and MR-RD model types, it is seen that one first-order feature, kurtosis, is high on the importance scale. Statistically, kurtosis is used to describe the sharpness of the distribution. A study by Wang et al.^38^ discusses the potential kurtosis has for quantifying tumor heterogeneity and the progression of lung disease. Hence, it would be of interest to study the relationship between the occurrence of RP and kurtosis to better understand how this feature plays a role in predicting RP. In all the multiomic models, it was the dosiomic features that were dominant emphasizing the need to have dosiomic features in OPMs when predicting RP.

There are a few limitations to our study. This study is a single institution, retrospective analysis. For this study, we have only used the original pretreatment CT and dose images for extracting features. There are studies that have examined the utility of various pre-processed (filtered) CT images, in addition to original CT image, to predict RP.^39,40^ A future scope of this work could be to test if features extracted from filtered images help in enhancing the models based out of original omic features; this is motivated by Demircioglu et al. whose study suggests that preprocessing filters can influence radiomic analysis, and they can improve predictive performance of models.^41^ Another limitation of this study is that we only use pretreatment CT for our omic analysis. A future scope of this study could be to determine if we can further enhance the performance for dosiomic-based toxicity models by introducing pretreatment PET radiomic features into our modeling scheme, similar to a study by Anthony et al.^42^ Due to the inclusion of many radiomic and dosiomic features in our analysis, a feature reduction technique was used to avoid overfitting, thereby potentially excluding features that could be having predictive value. However, given the data-driven nature of our feature reduction process, we were able to find predictive features. In addition to radiomics and dosiomics, including other omic data, such as genomics and proteomics, could also provide insightful information and as a future scope, they could be introduced into the modeling scheme to improve model robustness. An efficient and effective way of utilizing these features during the treatment planning phase remains to be one goal of our future study.

## Conclusion

In this work, we discovered that multiomic models are more efficient than DVH-only models at predicting ≥grade 2 RP. The best performing OPM included both whole-lung and multi-region multiomic features, thereby demonstrating that a restricted region extracted proximal to the GTV has utility for improving RP-based OPMs. Dosiomic features were the most important in all multiomic RP toxicity model types, thereby demonstrating that these features add significant value to predictive models of RP toxicity. Future outcome prediction models of radiation pneumonitis should include both multi-region multiomic and dosiomic features for the highest performing OPMs.

## Data Availability

Research data are stored in an institutional repository and anonymized data will be shared, following a 12 month embargo after the date of publication, upon request to the corresponding author once a data transfer agreement has been reached between the requestors institution and MD Anderson Cancer Center.

