## Supplementary Materials for "Multi-region Multiomic Random Forest Toxicity Modeling of Radiation Pneumonitis"

### Supplementary Material

**Supplementary Table S1**. Patient clinical and pathological characteristics grouped according to radiation pneumonitis severity (symptomatic RP, Grade ≥2; asymptomatic RP, Grade <2).

| Characteristic | | Total | RP < 2 | RP ≥ 2 | p-value |
|---|---|---|---|---|---|
| Patients (n) | | 329 | 244 | 85 | |
| Age (yrs) | | | | | |
| | Mean | 65.6 (33-88) | | | 0.77 |
| Gender | | | | | |
| | Male | 183 | 138 | 45 | 0.705 |
| | Female | 146 | 106 | 40 | |
| Smoking History | | | | | |
| | Non-smoker | 36 | 23 | 13 | 0.181 |
| | Prior smoker | 222 | 170 | 52 | |
| | Active smoker | 71 | 51 | 20 | |
| Karnofsky Score | | | | | |
| | <=60 | 13 | 10 | 3 | 0.45 |
| | 70-80 | 219 | 160 | 59 | |
| | 90-100 | 97 | 74 | 23 | |
| T Stage | | | | | |
| | T0/T1 | 66 | 49 | 17 | 0.41 |
| | T2 | 124 | 91 | 33 | |
| | T3 | 63 | 44 | 19 | |
| | T4 | 76 | 60 | 16 | |
| N Stage | | | | | |
| | N0 | 43 | 38 | 5 | 0.039 |
| | N1 | 40 | 31 | 9 | |
| | N2 | 172 | 127 | 45 | |
| | N3 | 74 | 48 | 26 | |
| M Stage | | | | | |
| | M0 | 317 | 234 | 83 | 0.55 |
| | M1 | 12 | 10 | 2 | |
| Histology | | | | | |
| | Adeno | 182 | 131 | 51 | 0.51 |
| | Squamous | 111 | 86 | 25 | |
| | Poorly differentiated NSCLC | 1 | 1 | 0 | |
| | NSC NOS | 17 | 11 | 6 | |
| | Mixed NSC | 1 | 0 | 1 | |
| | Large Cell | 3 | 3 | 0 | |
| | Carcinoid | 3 | 2 | 1 | |
| | Other | 8 | 7 | 1 | |
| | Unknown | 3 | 3 | 0 | |
| Induction Chemo | | | | | |
| | Yes | 42 | 30 | 12 | 0.70 |
| | No | 287 | 214 | 73 | |
| Concurrent Chemo | | | | | |
| | Yes | 324 | 239 | 85 | 0.33 |
| | No | 5 | 5 | 0 | |
| Tumor Location | | | | | |
| | RUL | 131 | 92 | 39 | 0.165 |
| | RML | 48 | 35 | 13 | |
| | RLL | 40 | 29 | 11 | |
| | LUL | 69 | 57 | 12 | |
| | LLL | 32 | 22 | 10 | |
| | Mediastinum | 9 | 9 | 0 | |
| GTV Volume | | | | | |
| | Mean | 129.57 (0 - 2017.15) | 123.55 | 146.67 | <0.001 |

*Abbreviations: n, number of patients; age, years; T Stage, tumor size; N Stage, number of nearby lymph nodes that have cancer; M Stage, metastasis status; Chemo, chemotherapy; GTV, gross tumor volume; NSCLC, non-small cell lung cancer; NOS, not otherwise specified; RUL, right upper lung; RML, right middle lung; RLL, right lower lung; LUL, left upper lung; LLL, left lower lung.*

**Supplementary Table S2**. DVH features grouped according to radiation pneumonitis severity (symptomatic RP, Grade ≥2; asymptomatic RP, Grade <2).

| DVH Metrics | | Total | RP < 2 | RP ≥ 2 | p-value |
|---|---|---|---|---|---|
| Mean Lung Dose (Gy) | | | | | |
| | Mean | 15.58 (2.46 - 23.41) | 14.47 | 18.73 | <0.001 |
| Max Lung Dose (Gy) | | | | | |
| | Mean | 77.90 (53.74 - 94.86) | 77.94 | 77.78 | 0.88 |
| rV5 (Gy) | | | | | |
| | Mean | 45.27 (8.92 - 89.29) | 43.23 | 51.06 | <0.001 |
| rV10 (Gy) | | | | | |
| | Mean | 35.13 (5.38 - 66.64) | 33.23 | 40.52 | <0.001 |
| rV15 (Gy) | | | | | |
| | Mean | 29.96 (3.73 - 49.90) | 28.09 | 35.25 | <0.001 |
| rV20 (Gy) | | | | | |
| | Mean | 26.30 (2.82 - 42.28) | 24.50 | 31.39 | <0.001 |
| rV25 (Gy) | | | | | |
| | Mean | 23.51 (1.91 - 39.53) | 21.69 | 28.67 | <0.001 |
| rV30 (Gy) | | | | | |
| | Mean | 21.13 (1.01 - 36.59) | 19.30 | 26.31 | <0.001 |
| rV35 (Gy) | | | | | |
| | Mean | 18.93 (0.55 - 33.31) | 17.14 | 24.01 | <0.001 |
| rV40 (Gy) | | | | | |
| | Mean | 16.87 (0.28 - 31.26) | 15.14 | 21.78 | <0.001 |
| rV45 (Gy) | | | | | |
| | Mean | 14.94 (0.008 - 29.12) | 13.31 | 19.55 | <0.001 |
| rV50 (Gy) | | | | | |
| | Mean | 13.05 (0.006 - 26.23) | 11.59 | 17.21 | <0.001 |
| rV55 (Gy) | | | | | |
| | Mean | 11.13 (0.00 - 24.24) | 9.87 | 14.69 | <0.001 |
| rV60 (Gy) | | | | | |
| | Mean | 9.01 (0.00 - 22.49) | 8.02 | 11.83 | <0.001 |
| RT Technique | | | | | |
| | IMRT | 191 | 145 | 46 | 0.01 |
| | IMPT | 47 | 40 | 7 | |
| | PSPT | 92 | 59 | 33 | |
| Fractions | | | | | |
| | Mean | 33.83 (15-60) | 33.74 | 34.1 | 0.99 |
| Rx Dose (Gy) | | | | | |
| | Mean | 69.43(45-120) | 69.54 | 69.14 | 0.19 |

*Abbreviations: rV, percent volume of structure receiving a given dose; IMRT, intensity-modulated radiation therapy; IMPT, intensity-modulated proton therapy; PSPT, passive scattering proton therapy; Rx, prescribed dose; Gy, gray.*

**Supplementary Table S3**. Results of model type performance on the training set, reported as mean of each metric with one standard deviation as the error along. Model type comparison with the base model type (DVH metric) for WL-RD, MR-RD, and MR-DVHRD models is conducted with a one-sided Wilcoxon signed-rank test.

| Model Name | Accuracy | AUC | AUPRC | Precision | Recall |
|---|---|---|---|---|---|
| **Base (DVH)** | 0.90±0.01 | 0.93±0.01 | 0.88±0.02 | 0.86±0.05 | 0.81±0.05 |
| **WL-RD** | 0.91±0.01*** | 0.95±0.01*** | 0.91±0.02*** | 0.90±0.04*** | 0.82±0.03*** |
| **MR-RD** | 0.97±0.00*** | 0.99±0.00*** | 0.98±0.00*** | 0.96±0.02*** | 0.95±0.03*** |
| **MR-DVHRD** | 0.96±0.01*** | 0.98±0.00*** | 0.97±0.01*** | 0.97±0.03*** | 0.91±0.03*** |

*p<0.05; **p<0.001; ***p<0.00001 – Significance level as compared to the base model

*Abbreviations: DVH, dose volume histogram; AUC, area under the receiver operating characteristic curve; AUPRC, area under the precision-recall curve; WL, whole lung; RD, radiomic & dosiomic; MR, multi-region.*

#### Univariate Analysis

The endpoint of this study was grade ≥2 RP. Demographic and clinical factors (gender, age, smoking status, stage, histology, chemotherapy type, tumor location, GTV volume, and performance status) were compared between patients who had RP <2 and RP ≥2, in addition to mean lung dose, max lung dose, and DVH metrics. Mann-Whitney U-test and Fisher's exact test were used for continuous and categorical variables, respectively, and a p-value <0.05 was considered statistically significant.

##### Radiomic and Dosiomic Feature List

##### Types of features – Shape, First Order and Second Order

Below is a list of all multiomic features analyzed in the study, along with a description of each feature.

I. **Shape-Based Features –** Derived from geometric properties of the whole lung and ring respectively.

1. Mesh Volume – Volume of the total ROI.
2. Voxel Volume – Volume of the voxel.
3. Surface Area – Area of the ROI.
4. Surface to Volume Ratio – Ratio between area and volume.
5. Sphericity – Measure of roundness of shape of tumor relative to sphere.
6. Maximum 2D diameter (Slice) – largest pairwise Euclidean distance between tumor surface mesh vertices in the row- column plane.
7. Maximum 2D diameter (Column) – largest pairwise Euclidean distance between tumor surface mesh vertices in the row- slice plane.
8. Maximum 2D diameter (Row) – largest pairwise Euclidean distance between tumor. surface mesh vertices in the column- slice plane.
9. Major Axis Length – Yields largest axis length of ROI-enclosing ellipsoid.
10. Minor Axis Length – Yields second largest axis length of ROI-enclosing ellipsoid.
11. Lease Axis Length – Yields smallest axis length of ROI-enclosing ellipsoid.
12. Elongation – Shows relationship between two largest principal components in ROI.
13. Flatness - Shows relationship between largest and smallest principal components in ROI.
14. Maximum 3D diameter - largest pairwise Euclidean distance between tumor surface mesh vertices.

II. **First Order Features –** Used to describe the distribution of values of individual voxels. They are histogram based and provide single voxel values for every feature.

1. Energy – Measure of magnitude of voxel values in an image.
2. Total Energy – Value of energy scaled by the volume of voxel in cubic mm.
3. Entropy – Randomness in image values.
4. Minimum – Least gray level intensity inside the ROI.
5. $10^{th}$ percentile – $10^{th}$ percentile of voxels included in the ROI.

6. $90^{th}$ percentile- $90^{th}$ percentile of voxels included in the ROI.
7. Maximum – Max gray level intensity within ROI.
8. Mean – Average gray level intensity within ROI.
9. Median – Median gray level intensity within ROI.
10. Interquartile Range – Interquartile range of gray level intensities within the ROI.
11. Range – Range of gray level intensities within the ROI.
12. Skewness – Measures the symmetry of the distribution of values about mean value.
13. Kurtosis – Peakedness of the distribution of values in the image ROI.
14. Variance – Mean of squared distances of each intensity value from Mean value.
15. Uniformity – Measure of sum of squares of each intensity value.
16. Mean Absolute Deviation – Mean distance of all intensity values from Mean value of image array.
17. Robust Mean Absolute Deviation – Mean distance of all intensity values from Mean Value calculated on the subset of image array with gray levels in between, or equal to the $10^{th}$ and $90^{th}$ percentile.
18. Root Mean Squared – Square root of the mean of squared intensity values.

III. **Second Order Features -** Known as texture features, describe statistical interrelationships between voxels. The texture features used in our study are derived from 4 texture-characterization matrices (Gray-Level Cooccurrence Matrix [GLCM], Gray-Level Dependence Matrix [GLDM], Gray-Level Run Length Matrix [GLRLM], Gray-Level Size Zone Matrix [GLSZM] and Neighboring Gray Tone Difference Matrix [NGTDM]).

**A. GLCM (Gray Level Co-occurrence Matrix) Features**

Describes the second-order joint probability function of an image region constrained by the mask. Features include -

i. Autocorrelation – Measure of magnitude of fineness and coarseness of texture.

ii. Joint Average – Mean gray level intensity of distribution.

iii. Cluster Prominence – Measure of skewness and asymmetry of GLCM.

iv. Cluster Shade - Measure of skewness and uniformity of GLCM.

v. Cluster Tendency – Measure of groupings of voxels with similar gray levels.

vi. Contrast – Measure of local intensity variation.

vii. Correlation - value between 0 (uncorrelated) and 1 (perfectly correlated) showing the linear dependency of gray level values to their respective voxels in the GLCM.

viii. Difference Average - measures the relationship between occurrences of pairs with similar intensity values and occurrences of pairs with differing intensity values.

ix. Difference Entropy - Measure of the randomness/variability in neighborhood intensity value differences.

x. Difference Variance - Measure of heterogeneity that places higher weights on differing intensity level pairs that deviate more from the mean.

xi. Joint Energy – Measure of homogeneous patterns in the image.

xii. Joint Entropy - measure of the randomness/variability in neighborhood intensity values.

xiii. Informational Measure of Correlation (IMC) 1 - assesses the correlation between the probability distributions of i and j (quantifying the complexity of the texture), using mutual information.

xiv. Informational Measure of Correlation (IMC) 2 - assesses the correlation between the probability distributions of i and j (quantifying the complexity of the texture), using mutual information.

xv. Inverse Difference Moment – It is a measure of the local homogeneity of an image

xvi. Inverse Difference Moment Normalized - measure of the local homogeneity of an image.

xvii. Inverse Difference - Another measure of the local homogeneity of an image.

xviii. Inverse Difference Normalized - Another measure of the local homogeneity of an image.

xix. Inverse Variance – Measures the inverse variance.

xx. Maximum Probability - occurrences of the most predominant pair of neighboring intensity values.

xxi. Sum Entropy - Sum of neighborhood intensity value differences.

xxii. Sum of Squares - Measure in the distribution of neighboring intensity level pairs about the mean intensity level in the GLCM.

**B. Gray Level Size Zone Matrix (GLSZM) Features**

Quantifies gray level zones in an image. A gray level zone is defined as the number of connected voxels that share the same gray level intensity. Features include -

i. Small Area Emphasis - Measure of the distribution of small size zones, with a greater value indicative of more smaller size zones and more fine textures.

ii. Large Area Emphasis - Measure of the distribution of large area size zones, with a greater value indicative of more larger size zones and more coarse textures.

iii. Gray Level Non-Uniformity - measures the variability of gray-level intensity values in the image, with a lower value indicating more homogeneity in intensity values.

iv. Gray Level Non-Uniformity Normalized - Measures the variability of gray-level intensity values in the image, with a lower value indicating a greater similarity in intensity values.

v. Size-Zone Non-Uniformity - Measures the variability of size zone volumes in the image, with a lower value indicating more homogeneity in size zone volumes.

vi. Size-Zone Non-Uniformity Normalized - Measures the variability of size zone volumes throughout the image.

vii. Zone Percentage - Measures the coarseness of the texture by taking the ratio of number of zones and number of voxels in the ROI.

viii. Gray Level Variance - Measures the variance in gray level intensities for the zones.

ix. Zone Variance - Measures the variance in zone size volumes for the zones.

x. Zone Entropy - Measures the uncertainty/randomness in the distribution of zone sizes and gray levels.

xi. Low Gray Level Zone Emphasis - Measures the distribution of lower gray-level size zones, with a higher value indicating a greater proportion of lower gray-level values and size zones in the image.

xii. High Gray Level Zone Emphasis - Measures the distribution of the higher gray-level values, with a higher value indicating a greater proportion of higher gray-level values and size zones in the image.

xiii. Small Area Low Gray Level Emphasis - Measures the proportion in the image of the joint distribution of smaller size zones with lower gray-level values.

xiv. Small Area High Gray Level Emphasis - Measures the proportion in the image of the joint distribution of smaller size zones with higher gray-level values.

xv. Large Area High Gray Level Emphasis - Measures the proportion in the image of the joint distribution of larger size zones with higher gray-level values.

xvi. Large Area Low Gray Level Emphasis - Measures the proportion in the image of the joint distribution of larger size zones with lower gray-level values.

**C. Gray Level Run Length Matrix (GLRLM) Features**

Quantifies gray level runs, which are defined as the length in number of pixels, of consecutive pixels that have the same gray level value. Features include -

i. Short Run Emphasis - Measure of the distribution of short run lengths, with a greater value indicative of shorter run lengths and more fine textural textures.

ii. Long Run Emphasis - Measure of the distribution of long run lengths, with a greater value indicative of longer run lengths and more coarse structural textures.

iii. Gray Level Non-Uniformity - Measures the similarity of gray-level intensity values in the image, where a lower GLN value correlates with a greater similarity in intensity values.

iv. Gray Level Non-Uniformity Normalized - Measures the similarity of gray-level intensity values in the image, where a lower GLNN value correlates with a greater similarity in intensity values.

v. Run Length Non-Uniformity - Measures the similarity of run lengths throughout the image, with a lower value indicating more homogeneity among run lengths in the image.

vi. Run Length Non-Uniformity Normalized - Measures the similarity of run lengths throughout the image, with a lower value indicating more homogeneity among run lengths in the image.

vii. Run Percentage - Measures the coarseness of the texture by taking the ratio of number of runs and number of voxels in the ROI.

viii. Gray Level Variance - Measures the variance in gray level intensity for the runs.

ix. Run Variance - Measure of the variance in runs for the run lengths.

x. Run Entropy - Measures the uncertainty/randomness in the distribution of run lengths and gray levels. A higher value indicates more heterogeneity in the texture patterns.

xi. Low Gray Level Run Emphasis - Measures the distribution of low gray-level values, with a higher value indicating a greater concentration of low gray-level values in the image.

xii. High Gray Level Run Emphasis - Measures the distribution of the higher gray-level values, with a higher value indicating a greater concentration of high gray-level values in the image.

xiii. Short Run Low Gray Level Emphasis - Measures the joint distribution of shorter run lengths with lower gray-level values.

xiv. Short Run High Gray Level Emphasis - Measures the joint distribution of shorter run lengths with higher gray-level values.

xv. Long Run Low Gray Level Emphasis - Measures the joint distribution of long run lengths with lower gray-level values.

xvi. Long Run High Gray Level Emphasis - Measures the joint distribution of long run lengths with higher gray-level values.

**D. Neighboring Gray Tone Difference Matrix (NGTDM) Features**

Quantifies the difference between a gray value and the average gray value of its neighbors within distance “d”. Features include -

i. Coarseness - Measure of average difference between the center voxel and its neighborhood and is an indication of the spatial rate of change.

ii. Contrast - Measure of the spatial intensity change but is also dependent on the overall gray level dynamic range.

iii. Busyness - Measure of the change from a pixel to its neighbor.

iv. Complexity - An image is considered complex when there are many primitive components in the image, i.e. the image is non-uniform and there are many rapid changes in gray level intensity.

v. Strength - Measure of the primitives in an image.

**E. Gray Level Dependence Matrix (GLDM) Features**

Quantifies gray level dependencies in an image. Features include -

i. Small Dependence Emphasis - Measure of the distribution of small dependencies, with a greater value indicative of smaller dependence and less homogeneous textures.

ii. Large Dependence Emphasis - Measure of the distribution of large dependencies, with a greater value indicative of larger dependence and more homogeneous textures.

iii. Gray Level Non-Uniformity - Measures the similarity of gray-level intensity values in the image, where a lower GLN value correlates with a greater similarity in intensity values.

iv. Dependence Non-Uniformity - Measures the similarity of dependence throughout the image, with a lower value indicating more homogeneity among dependencies in the image.

v. Dependence Non-Uniformity Normalized - Measures the similarity of dependence throughout the image, with a lower value indicating more homogeneity among dependencies in the image.

vi. Gray Level Variance – Measures variance in gray level in the image

vii. Dependence Variance - Measures the variance in dependence size in the image.

viii. Dependence Entropy – Measures Entropy in the image

ix. Low Gray Level Emphasis - Measures the distribution of low gray-level values, with a higher value indicating a greater concentration of low gray-level values in the image.

x. High Gray Level Emphasis - Measures the distribution of the higher gray-level values, with a higher value indicating a greater concentration of high gray-level values in the image.

xi. Small Dependence Low Gray Level Emphasis - Measures the joint distribution of small dependence with lower gray-level values.

xii. Small Dependence High Gray Level Emphasis - Measures the joint distribution of small dependence with higher gray-level values.

xiii. Large Dependence High Gray Level Emphasis - Measures the joint distribution of large dependence with higher gray-level values.
